# Integrating psychosocial variables and societal diversity in epidemic models for predicting COVID-19 transmission dynamics

**DOI:** 10.1101/2020.08.12.20173252

**Authors:** VK Jirsa, S Petkoski, H Wang, M Woodman, J Fousek, C Betsch, L Felgendreff, R Böhm, L Lilleholt, I Zettler, SM Faber, K Shen, AR McIntosh

## Abstract

During the current COVID-19 pandemic, governments must make decisions based on a variety of information including estimations of infection spread, health care capacity, economic and psychosocial considerations. The disparate validity of current short-term forecasts of these factors is a major challenge to governments. By causally linking an established epidemiological spread model with dynamically evolving psychosocial variables, using Bayesian inference we estimate the strength and direction of these interactions for German and Danish data of disease spread, human mobility, and psychosocial factors based on the serial cross-sectional COVID-19 Snapshot Monitoring (COSMO; *N* = 16,981). We demonstrate that the strength of cumulative influence of psychosocial variables on infection rates is of a similar magnitude as the influence of physical distancing. We further show that the efficacy of political interventions to contain the disease strongly depends on societal diversity, in particular group-specific sensitivity to affective risk perception. As a consequence, the model may assist in quantifying the effect and timing of interventions, forecasting future scenarios, and differentiating the impact on diverse groups as a function of their societal organization. Importantly, the careful handling of societal factors, including support to the more vulnerable groups, adds another direct instrument to the battery of political interventions fighting epidemic spread.

## Introduction

The current Coronavirus disease 2019 (COVID-19) pandemic is more than a health crisis; it is also an economic, social and, in many countries, political crisis. Governmental decisions to fight COVID-19 must consider a range of factors including infection spread, health care capacity, as well as economic and psychological variables, all embedded and linked in a worldwide dynamic context. Importantly, these variables influence each other and evolve over time. The specific trajectory of these interactions will determine the consequences for each nation (e.g., deaths, duration of lock-down, economic loss, etc.). Indeed, for the battle against COVID-19 to be successful, it has been suggested to “inform and qualify action with evidence from behavioral and cultural research” (*1*) to create high acceptance of and compliance with measures, even after restrictions are lifted. While science offers this evidence to inform policy making (*2,3*), the influence of psychosocial (PS) variables is not integrated into epidemiological models to predict further disease transmission. Feeling risk of contracting the disease but also other, more remote psychosocial constructs such as trust in the regulating bodies and acceptance of policies (*1*) may be important to take up protective behavior (e.g., physical distancing, wearing a face mask). In current approaches the time invariance of these behaviors is assumed and adapted when an intervention occurs (*4*). Any violation of model assumptions reduces the predictive power of the model and limits its validity to short terms. A potential interaction of psychosocial variables with mobility is such a violation: high psychological strain felt during a lock-down may lead people to reject the preventative measures, which is then related to increased mobility (*5*). Consequently, model parameters need to be continuously updated to counter for the accumulating model errors (*6, 7*). As governmental decisions and interventions need to be planned further ahead, the mechanistic correction of the forecast model by integrating the interactions of PS variables with mobility is crucial. Here we use the weekly cross-sectional COVID-19 Snapshot Monitoring (COSMO) data (*5*, *8*), which allow rapid and adaptive monitoring of psychosocial variables and gain a real-time insight into the knowledge, perceptions, and behaviors of the population – the “psychological COVID-19 situation” (*9*). Our objective is to improve the accuracy of longer-term predictions of epidemic spread models by integrating the influence of PS variables into disease dynamics.

## Integrating psychosocial variables in epidemic transmission model of COVID-19

Studies such as COSMO suggest that as PS variables change over time, behavioral responses will change over a similar time scale. The temporal alignment of PS variables, mobility, and infectious spread provides a framework that enables a causal analysis and testing of this hypothesis (*10*). As a first step, we express the influence of PS variables upon behavior as a causal hypothesis, where its coupling strength and direction is inferred from the empirical data. Changes in behavior are expressed by modulation of the contact rate and quantified by the mixing matrix *β*, which describes the number of infection-producing contacts per unit time. R_0_ is the reproduction number of an infection and reflects the expected number of cases directly generated by one case in a homogeneously mixed population where all individuals are susceptible to infection (*11*). Both quantities, *β* and R_0_, cannot currently be modified through vaccination or other changes in population susceptibility, but can be modified by physical distancing and other political interventions. These measures, however, are only effective if large parts of the population comply with them. However, certain groups in the population may have less resources and capacity to adapt and thus perceive different levels of affect, e.g. due to age or variations in personal situations, such as a loss of income during a lock-down, exposed working conditions, or restricted living space. We hypothesize that these groups will show different degrees of acceptance of the measures, leading to variation in mobility and ultimately disease transmission, which then again causally loops onto itself, contributing to further changes in PS levels.

We formalize this emergent and dynamic process in Figure 1, using a compartmental SIR model for infectious disease transmission comprising three populations of susceptible (S) and infected (I) cases, as well as recovered or dead (R) cases. The total number of people is N=S+I+R. As a government imposes political interventions *J* such as a lock-down, the mixing matrix *β* and the subsequent reproduction number R_0_ are reduced. Additional seasonal modulations of *β* add an important temporal variation to the epidemic transmission mechanisms (*12*). We expand *β* along two additional dimensions. First, *β* can slowly evolve over time as a consequence of modulations with the PS variables. The time scale separation between infectious transmission and PS variables justifies the mathematical assumption of weak coupling and its expression as an additive modulation (*13*). Second, groups within the population can be differentially affected depending on their resources and capacity to adapt to the situation (1), which we separate into *vulnerable* and *resilient* groups for the purposes of the model. The affective state *ϕ_i_* of the *i*-th group is a longer lasting state, which is not caused by a single stimulus but results from an accumulation of experiences. The accumulation of PS effects, *u_i_*, accrues over time due to prolonged restrictions, indicating the capacity of the *i-th* group to adapt to the imposed political interventions. It varies with the pandemic containment efforts such as physical distancing and travel restrictions, but also economic factors such as business closures. Behavioral changes are absorbed in the mixing matrix *β_i_*, which establishes the link to the SIR variables and only depends on the political interventions J and the affective state *ϕ_i_* of the *i-th* group.

**Figure 1.**
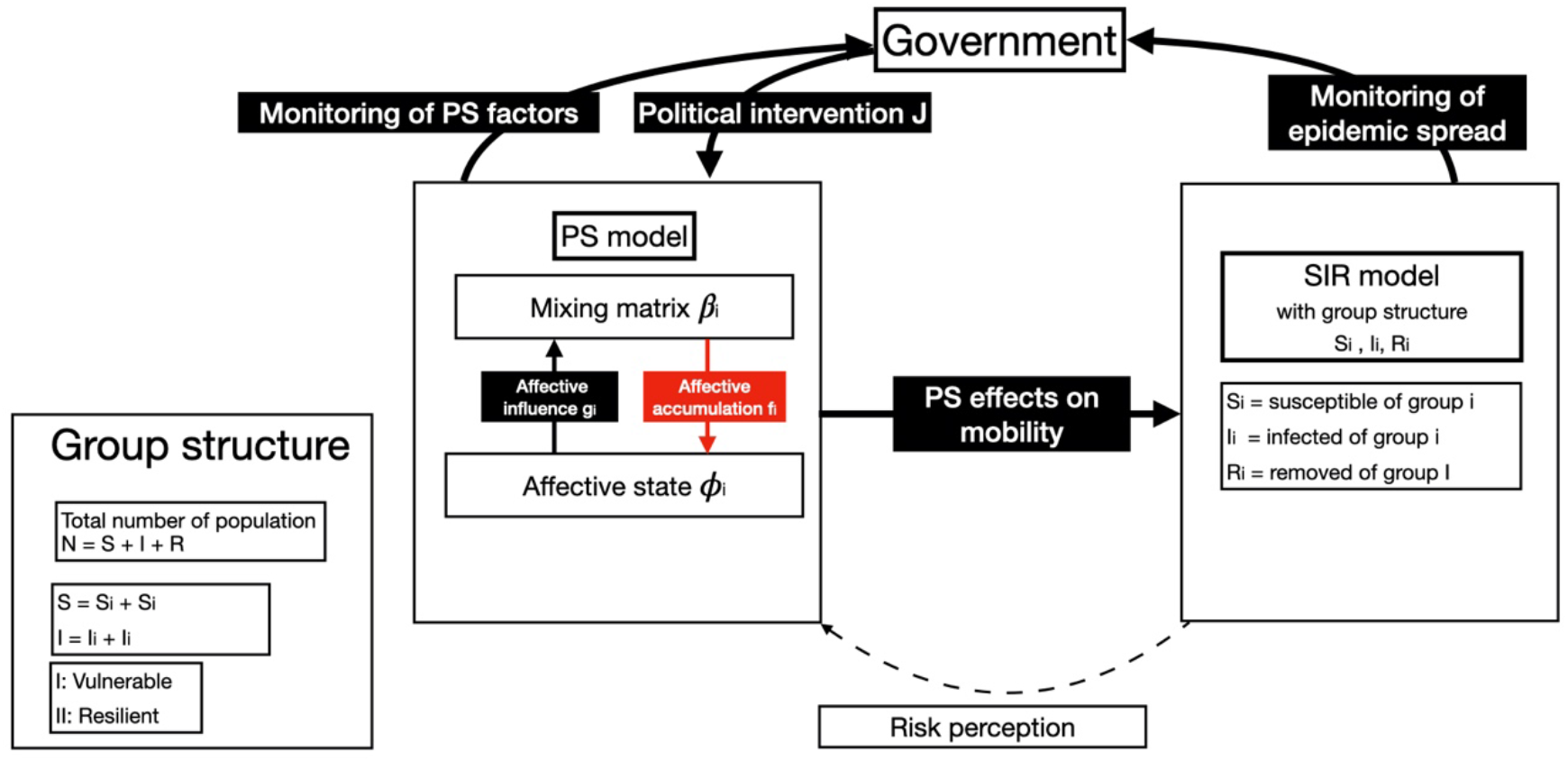
Organization of the PS-SIR model. Psychosocial (PS) variables modulate human mobility through changes of the mixing matrix *β_i_*. As the perceived consequences of political interventions, such as lock-down effects, accrue, the subsequent accumulation changes the affective state *ϕ_i_* of the i-th sub-population. *β_i_* affects *ϕ_i_* through the influence *f_i_* of accumulated PS variables and *ϕ_i_* affects *β_i_* through the influence *g_i_*, e.g., affective risk perception. The index i signals group-specific variation of parameters and variables across the whole population, defining a group structure of vulnerable vs. resilient groups with varying resources and capacity to adapt. Changes in the mixing matrix *β_i_* directly affect the dynamics of the infectious disease in the SIR model.

To infer the strength and direction of the interactions, we use the COSMO data of Germany and Denmark. The measures derived from the COSMO data sets were collected weekly from early-March in Germany (overall *N* = 11,669) and started a few weeks later in Denmark (overall *N* = 5,312). The measures focused mainly on the perception of the COVID-19 pandemic, including: AFFECT variables that estimated how serious the threat was to the respondent (e.g., “Is the virus near to you?”; “Is the virus spreading fast?”; “Are you worried about the virus?”), POLICY adjustments and acceptance (e.g., “Should the government close schools?”) and TRUST in institutions (“Do you trust the performance of the government?”; “Do you trust the healthcare system?”; see Appendix for more details on questions). Germany and Denmark show both an early increase in the concern around the spread of the virus (AFFECT), and agreement with the governmental interventions (POLICY), especially around school closings. This peaked around March 23, when the country-wide lock-down was instituted in Germany (Figure 2). This trend declined across time, indicating less concern with the pandemic, to levels similar to where they stood when data were first collected. The data from Denmark were collected starting immediately after the country-wide lock-down but showed a comparable declining trend with more recent measurement waves. Figure 2 shows the time-aligned COVID-19 infection and death rates over the same time period (*4*), and the mobility of the population based on cell phone tracking indicating sharply reduced mobility during the lockdown and then increasing mobility as AFFECT and POLICY declined (*14*).

**Figure 2.**
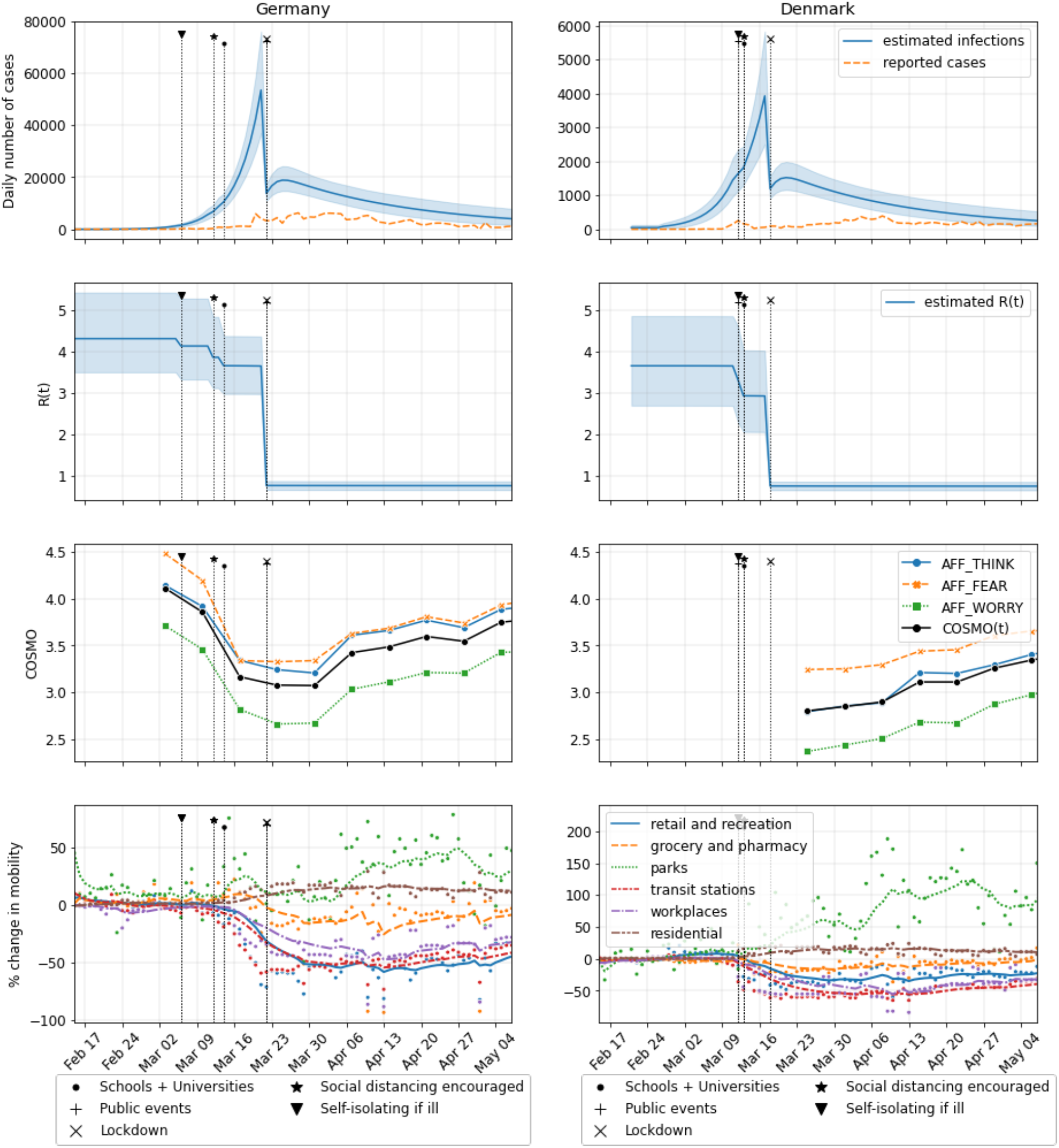
Data available for Germany and Denmark from February 15th to early of May 2020. In the first two rows, estimates for daily infections *(4)* and reproduction number R_0_ are presented as time varying means and standard deviations. Third row shows three selected variables from the AFFECT group in the COSMO surveys (see Appendix for more details; higher values indicate that COVID-19 was seen as something one does not think about all the time, rather not terrifying, and something not to worry about); their weighted mean (derived from PCA) labeled as COSMO(t) is used in the model inversion. The last row presents the scatter plots of the Google mobility report overlaid with lines marking the 7-day weighted moving averages, where the mean of the blue “retail and recreation” variable is used in the model inversion.

## Sampling of psychosocial and behavioral interactions during COVID-19

In order to assess the strength, direction, and significance of the coupling of PS variables on the time-varying reproduction number R_0_, we submit the above extended SIR model with PS interactions to Markov Chain Monte Carlo (MCMC) Bayesian inference. Three parameters are inferred: *γ* and *c* are the strength and direction of the couplings *g_i_* and *f_i_*, respectively (Figure 1), and *Γ* is the time scale of PS accumulation. For simplicity we dropped the group-specific indices of *γ* and *c*, as we do not have access to group-specific mobility data from Google. Figure 3 shows the results of the inference and the data used in the process. The two top rows show the time series for the reproduction number R_0_(t) estimates (*4*) for Germany (DE) and Denmark (DK), respectively, and the corresponding fit of the model as lines with the same color scheme. The bottom row shows posterior distributions for model parameters of interest along with the priors (in grey) placed on those parameters. While in Bayesian terms, the coupling strength *γ* is well identified by the data of both Denmark and Germany, *Γ,c* are identified strongly only for data from Germany. For Denmark these posteriors follow the prior, which are likely due to the lack of COSMO data during the onset of confinement in Denmark. As our primary goal is to determine whether *γ* is non-zero, i.e., PS factors have an effect on reproduction rate, we implement our null hypothesis as a normal prior on *γ*, centered at 0.1, i.e., a small effect. MCMC sampling from the posterior distribution allow the data to then identify *γ, Γ, c*. The point estimates (medians) of *γ* are 0.21 and 0.23 for Germany and Denmark, respectively, and 95% confidence intervals are (0.17, 0.25) and (0.18, 0.30) for Germany and Denmark, respectively. This indicates that PS variables significantly impact the reproduction rate with a non-zero and positive coupling strength, i.e. the less stress people felt about COVID-19, the higher R0. This defines a baseline, which allows individual parametrization of the models for Germany and Denmark.

**Figure 3:**
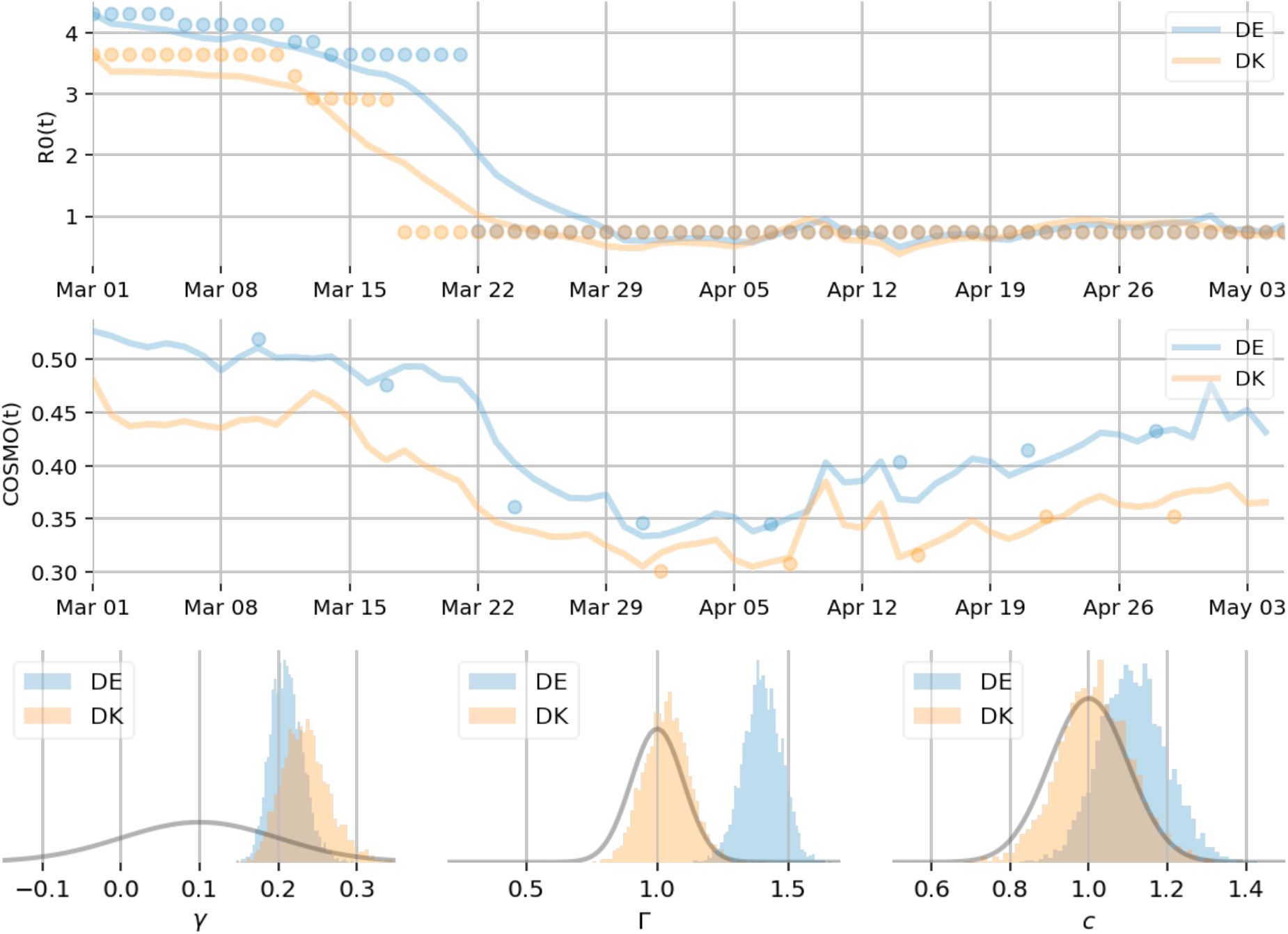
Inference of PS-SIR model parameters. *Top*: time series are shown for R_0_(t), time varying reproduction rate estimates (see Fig 2), for Germany (DE) and Denmark (DK) in blue and orange circles, respectively, and the fit of the model as lines with the same color scheme. *Middle:* time series are shown for COSMO AFFECT data, for Germany and Denmark in blue and orange circles, respectively, and the fit of the model as lines with the same color scheme. Lower values indicate higher negative affect such as more fear and worries due to the Coronavirus. *Bottom:* MCMC posterior distributions for key model parameters (*γ:* coupling strength of *g_i_*; *Γ*: time scale of psychosocial accumulation; *c*: coupling strength of *f_i_*) for Germany and Denmark (blue and orange histograms, respectively) and their respective priors (grey lines). On the left, the histograms of posterior samples for *γ* for both countries, which deviate significantly from the prior (grey line); the data thus identify a strong and robust effect of PS factors on SIR dynamics. Similar effects are seen for *Γ* and *c* for Germany, while Denmark follows the prior on these parameters likely because of missing COSMO data at the onset of confinement.

## Exploring the effects of PS variation on pandemic spread

We initialize each model with its respective past values for epidemic, mobility, and PS variables and provide simulations for the upcoming months. To estimate robustness of the forecast simulations and sensitivity upon the inferred parameters, we run simulations for all values of coupling parameters sampled from the posterior distributions. The panels of Figure 4 show a comparison of simulations for the scenarios with and without PS coupling, displaying the forecast trajectory evolution until April 2021 for the number of infected individuals *I*, reproduction number R_0_, affective state *ϕ_i_*, and estimated strength of political interventions *J*. The left column displays the situation for Germany, the right for Denmark. Further differentiation comprises group structure as indexed by *i*, simulating the population into groups of vulnerable and resilient based on different levels of PS variables in response to political interventions. The group differentiation is realized by assigning different values to coupling parameters *c_i_* and *Γ_i_*.

**Figure 4:**
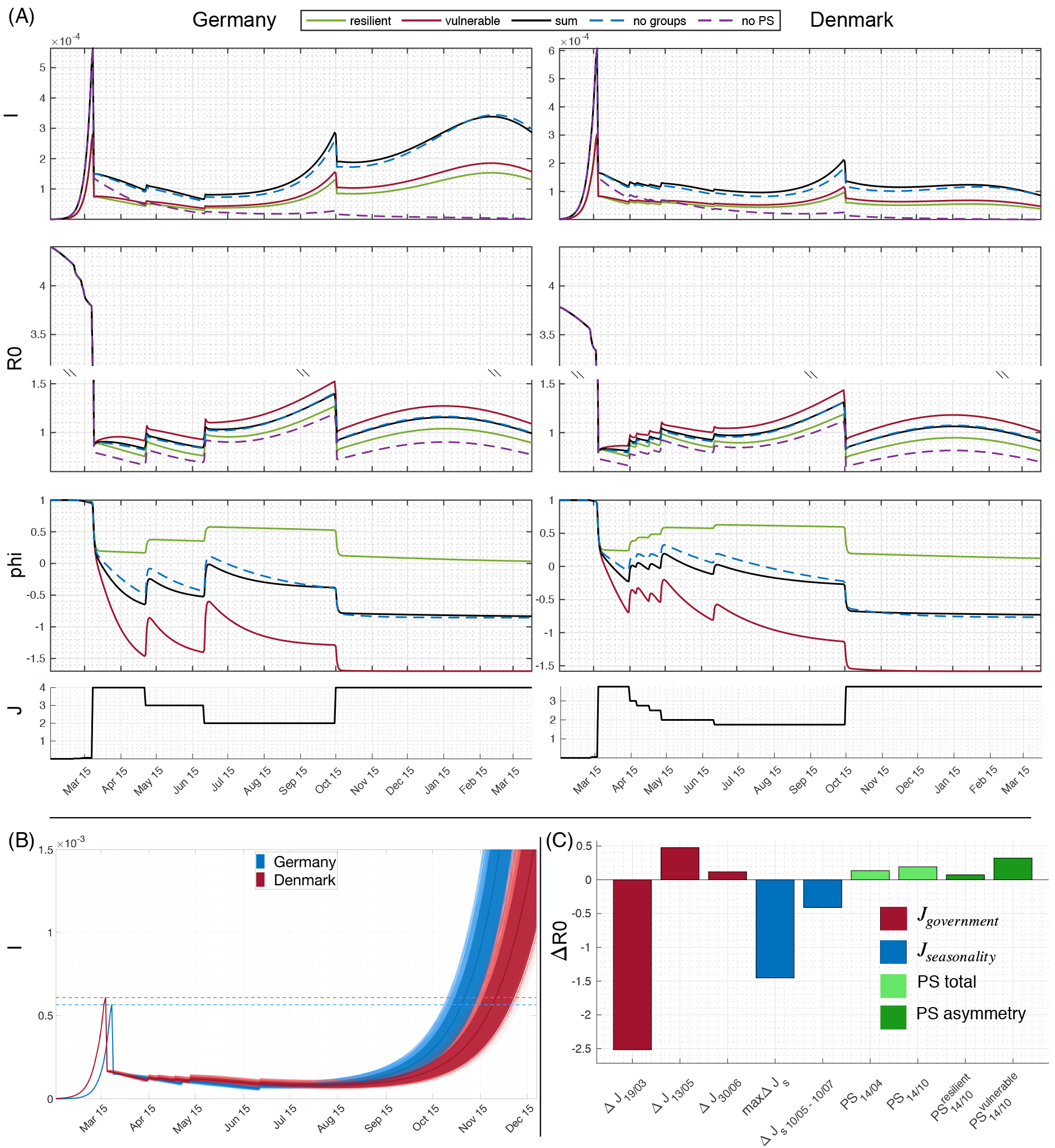
Forecast of the evolution of COVID 19 spread for Germany and Denmark. (A) Simulations of the SIR (dashed magenta), PS-SIR model with (black) and without group structure (dashed blue) for Germany (left) and Denmark (right) as well as the PS-SIR model split up for populations assuming 50% vulnerable (red) or resilient (green) citizens, simulated until April 2021. “I” denotes the number infected, R0 the reproduction factor, phi perceived negative AFFECT, and J political interventions. The default parameters in all the plots are from a run of the chain that best fits the data shown in Figure 2 up to mid-May. Simulations contain all the government interventions up to the end of June, as well as a re-imposing of the strictest previous intervention in early autumn in order to prevent the second infectious wave due to the seasonality. (B) Ensemble projection of infections for all the combinations of inferred parameters for both countries (light red and blue lines), the mean (red and blue line) and the areas of 95% confidence (shaded red and blue). (C) Estimated strengths of interventions on infectious spread for Denmark (results for Germany are comparable and provided in the Appendix, Figure S2): Red is the impact of the government interventions (the largest on 19/03, and the subsequent relaxations on 13/05 and 30/06); blue is seasonality (the maximum impact, and the impact for 2 months after the relaxation on 10/05 to 10/07). Green is the impact of the PS subsystem, where light green is the cumulative impact before the relaxation on 14/04 and before the predicted intervention in autumn; and dark green is the predicted impact in autumn 2020, separated per group.

The seasonality of transmission *(12)* is included in the simulations where more transmissions are assumed in autumn/winter than in spring/summer. Together, PS variables and seasonality cause the predicted epidemic wave in autumn/winter of 2020 to be larger than the previous one in spring 2020, despite the same stringency level of lock-down. This effect is also partly responsible (together with the government interventions) for the lower level of infections observed during May and June 2020 than later on in autumn. For every simulation of the SIR model without PS variables (magenta dashed lines in Figure 4A), the efficacy of political interventions *J* is overestimated. When comparing the forecast epidemic spread with and without PS coupling (*γ*=0), the political intervention *J* would have proven sufficient to control the epidemic spread in the latter case, but not the former, because the PS variables cause a surge of infections towards the second part of 2020. Even though the mean inferred impact of the PS subsystem is slightly lower for Germany than for Denmark, *R*_0_ and the second infection wave related to the seasonality is predicted to be larger in Germany. The sensitivity of the simulations to the choice of parameters is explored in Figure 4B. Here the coupling parameters *γ* are drawn from the posterior distributions for a sample of 4,000 model realizations for each country, and then used in the simulations with all other parameter settings equal. The shaded areas indicate a confidence interval of 95%. All simulated trajectories show qualitatively the same behavior, that is a surge of infection towards the end of 2020. To provide a quantitative impression of the strength of PS coupling as compared to lock-down and seasonal variation, Figure 4C shows a comparison of the impact of different interventions with the seasonality and the PS influence, illustrated for the case of Denmark. Analysis for Germany provides the same results and is provided in the appendix Fig S2. The inclusion of group structure into the PS coupling generally enhances its strength, where more heterogeneity cause stronger PS effects (see also appendix Fig S1 for further elaboration). The impact is computed as the difference of cumulative infections with and without cause (intervention, seasonality, and PS variables). Given the cumulative nature of PS effects, the effect strength has been inferred for a particular time after the onset of the cause, in particular the cumulative impact of PS is shown before the relaxation on 14/04 and before the predicted intervention in autumn (light green); furthermore, the cumulative impact of group structure is also shown (dark green), clearly demonstrating its epidemic spread-reinforcing effect. Political interventions via mobility restrictions show the strongest effect, seasonality is factor 2 smaller than mobility constraint, and a factor 3 to 4 larger than PS factors.

## Grouping PS variables in the COSMO data

Given the importance of the group structure on the relation between PS variables and SIR suggested by the model, we further analyzed the COSMO survey data to determine if significant grouping variables could be extracted. We tested age (young, middle, and old age), sex, and education (three levels) and compared the aggregate PS variables across all available waves using multivariate Partial Least Squares (PLS) analysis (*15, 16)*. Employment status and household income were less consistently collected so were not considered for this analysis. In both German and Danish data, differences based on sex (Figure 5) and age (Supplementary Figure S1) were observed. Differences related to education were less reliable but existent. We also observed that the aggregate PS measure was reliably related to subjective psychological stress and resilience in a subset of the COSMO data, suggesting the aggregate captured broader psychosocial aspects beyond risk perception (see Appendix Fig S4). For Germany, data collection started earlier and shows an initial overlapping transient with a faster rebound for males than for females (Figure 5 left), resulting in a group difference which remains preserved as time evolves. The same situation is observed for Denmark, even more pronounced (Figure 5 right), but without the initial transient as data collection started later. The interaction of age and sex was not significant, indicating these were additive factors in these data. Older adults and females showed larger overall values on the PS effect, suggesting these groups felt higher affective risk and that the COVID-19 pandemic was more of a crisis relative to younger groups or males.

**Figure 5.**
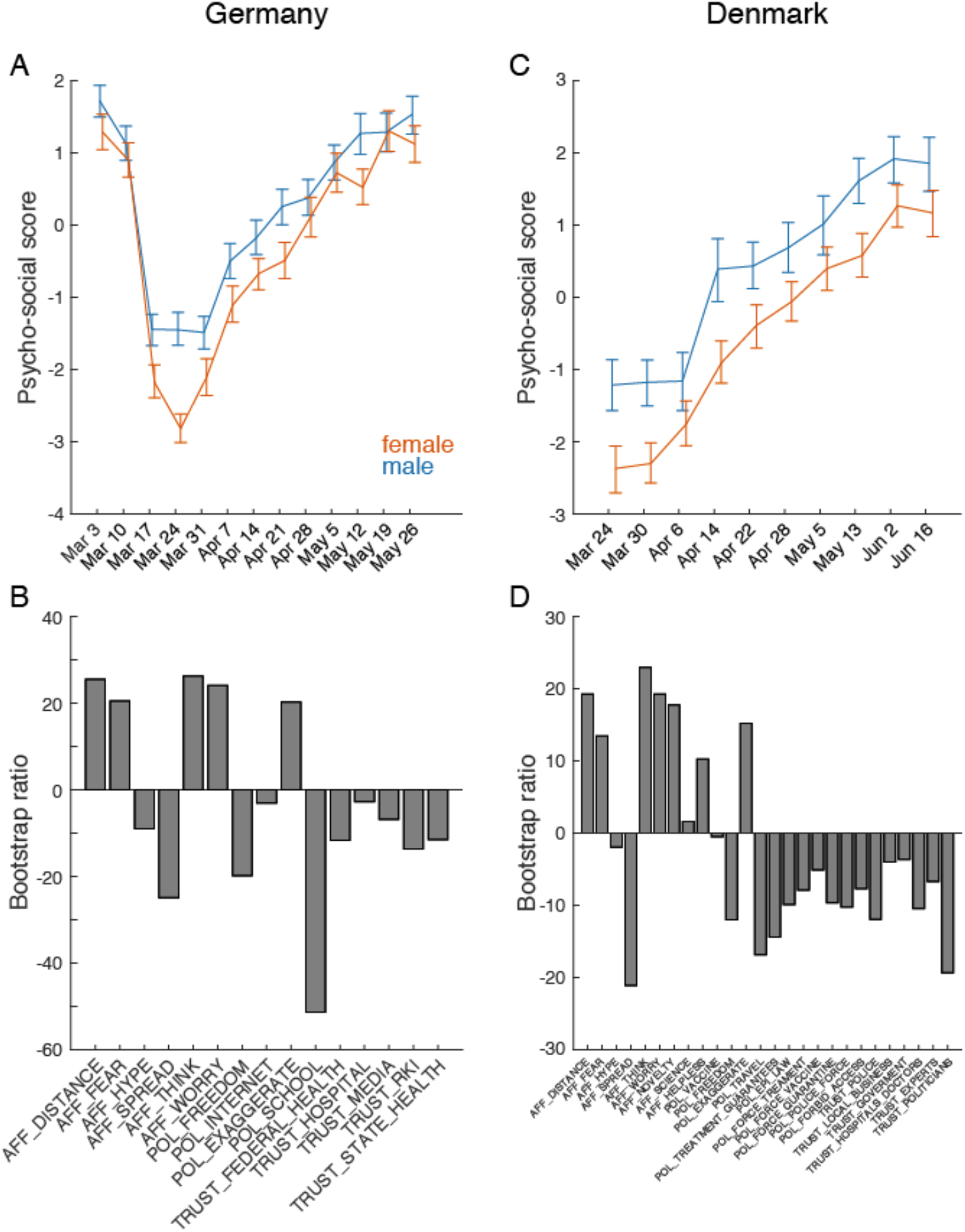
Sex differences in PS indicators. The dominant effect (both p < 0.001 from permutation test) of a multivariate Partial Least Squares analysis of mean change over time of PS indicators in COSMO data from Germany (A-B) and Denmark (C-D) by sex. AFFECT (AFF), POLICY (POL) and TRUST variables were included if data were available from all waves in each of the COSMO datasets. See Supplementary Table 1 for a complete list of variables. Error bars: Bootstrap estimated 95% CI. Bootstrap ratios are the singular value weights divided by their standard error and are roughly equivalent to a z-score. The shape of the Psychosocial score curve relative to the PS Indicators suggests increase in affect risk perception and great institutional and policy trust in March, followed by a general relative reduction.

## Discussion

Although mechanisms of COVID-19 transmission are indiscriminate, this may not extend to a complex society fighting epidemic spread *(17)*. Societies have a rich inter-individual variability, different cultures, wealth distribution, and ways of functioning, but most epidemic models describe the dynamics of a homogeneous population in different disease stages. The model parameters absorb all properties characterizing the diversity of society including psychosocial factors. With the introduction of psychosocial coupling in epidemic disease models, the consideration of effects linked to societal diversity becomes possible.

We estimated the strength and direction of psychosocial causes within a causal inference framework and demonstrated their effect upon the infectious spread to be smaller, but on a similar scale of magnitude as current political interventions (approx. factor 8 smaller than lockdown effects and social distancing) and seasonal variation (approx. factor 4-5 smaller). In particular, affect variables that estimated how serious the threat was to the respondent provide strong evidence for the causal effect of psychosocial factors on mobility and subsequently infectious spread. Our cause-effect analysis considered minor parametric changes due to psychosocial variation, but has the capacity to integrate a large range of multi-dimensional data including societal, national and cultural differences, political strategies, and trust in the government and media. For instance, multidimensional approaches could integrate additional psychological and sociocultural indicators, affecting the mixing matrix, thereby establishing a battery of mutually dependent behaviors. Given the here estimated size of psychosocial causes, the synergy of these interactions may likely reach scales that cannot be ignored in epidemic spread. Furthermore, heterogeneity within a society generates groups that are not equally affected by the same political intervention. The group structuring differences are plotted in Figure 5 and supplemental figures, demonstrating that a highly heterogeneous society would show acceleration and augmentation of infections across the entire population due to group structure (Figure S1). Despite the fact that the resilient population experiences no change in its affective state, the uniform mixing of the populations causes increases of total numbers of infections in both groups, vulnerable and resilient. A detailed quantitative understanding of these effects provides an added means to guide political decisions to influence the infectious spread by considering parameters linked to demographic factors of societal organization. In the continuum spanned by group heterogeneity and timing of interventions, the choice of the best lock-down duration will depend on the heterogeneity within the social structure and allow the optimization of political interventions for a given society, and thus minimize economic damage while respecting the efficiency for epidemic transmission control. This is particularly important as interventions have optimal working domains (such as lock-down strength or duration), but outside of these windows they are inefficient. For example, in Figure S1A for Denmark, showing that extending lock-down durations do not alter the trajectory of epidemic spread significantly. Alternatively, the same mechanism in the our model may be used to increase the efficacy of political interventions by changing societal factors, in particular reducing societal heterogeneity and increasing support to the more vulnerable groups (*1*).

Partly because of a lack of specific data in the COSMO surveys, we have left explicit economic factors aside in this discussion, although such factors would be relevant to the grouping structure. It has been suggested that psych-socio-economic variables such as income, education, and ethnicity play an important role in the transmission of infectious diseases (*18–20*), not only because they reflect resilience, but also because they influence the adoption of protective behaviors during pandemics (*21*). As a consequence, models taking into consideration PS-SIR interactions can provide stronger support for political interventions that support less resilient groups socially and economically as this is a factor in reducing heterogeneity (*22, 23)*. The model framework also underscores that psychological knowledge from personality and health psychology can be put to very concrete use in a crisis (*24*). Future efforts should quantitatively assess the impact of communication and behavioural interventions that address PS factors to enhance compliance.

The COVID-19 pandemic is exacerbating existing global and national inequalities, which typically hit the most vulnerable groups hardest *(25, 26)*. The focus of policymakers has been on containing the spread of COVID-19 and mitigating the socioeconomic effects of the pandemic. Our findings demonstrate that addressing these vulnerabilities nation- or even worldwide through policy action (for instance, temporary basic income strategy *(17, 27, 28)* may not only be a mean to fight the socioeconomic consequences of COVID-19, but also represent a mean to directly fight its spread across all groups, both vulnerable and resilient, through non-pharmaceutical actions containing the disease.

## Data Availability

All simulated data are available upon request.

https://projekte.uni-erfurt.de/cosmo2020/cosmo-analysis.html

## Acknowledgements

This research was supported by European Union’s Horizon 2020 research and innovation programme under grant agreement No. 945539 (SGA3) Human Brain Project. Germany’s COVID-19 Snapshot Monitoring (COSMO) is a joint project of the University of Erfurt (Cornelia Betsch [PI], Lars Korn, Philipp Sprengholz, Philipp Schmid, Lisa Felgendreff, Sarah Eitze), the Robert Koch Institute (RKI; Lothar H. Wieler, Patrick Schmich), the Federal Centre for Health Education (BZgA; Heidrun Thaiss, Freia De Bock), the Leibniz Centre for Psychological Information and Documentation (ZPID; Michael Bosnjak), the Science Media Center (SMC; Volker Stollorz), the Bernhard Nocht Institute for Tropical Medicine (BNITM; Michael Ramharter), and the Yale Institute for Global Health (Saad Omer); funding: BZgA, RKI, ZPID, University of Erfurt (no grant number). Denmark’s COSMO is funded by the Faculty of Social Sciences, University of Copenhagen (to RB and IZ), and Lundbeck Foundation, grant no. R349-2020-592 (to RB).

## Methods

### Model

We express the influence of *ϕ_i_* and *β_i_* by the coupling functions *f_i_(u_i_,J,ϕ_i_*) and *g_i_*(*ϕ_i_,J,β_i_*), which shall be inferred from the data. As a first step in model building, we recognize similarities with many physical and biological systems, which are coupled indirectly through a slowly evolving dynamic medium. Such slow modulations of a coupling medium are effectively equivalent to a linear weak coupling of the dynamic systems, prescribing their interaction around a working point as additive in a first approximation, but leaving the mathematical form of the actual interaction functions themselves open. We harness this effect to extend the transmission dynamics of the SIR model to include psychosocial interactions and infer the precise form of the influence functions using Monte Carlo Markov Chain (MCMC) methods. The full set of equations reads:

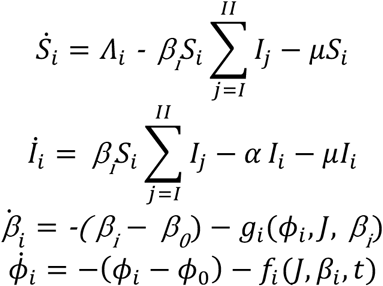

where the coupling functions *g_i_*(*ϕ_i_,J, β_i_*) and *f_i_ (J,β_i_*, t) affect the growth rates of the mixing matrix *β_i_* and affective state *ϕ_i_*. The back-coupling of *β_i_* on *ϕ_i_* is communicated via an accumulation of lock-down effects, aka deviations from normal mobility, through an auxiliary variable *u_i_*. For both types of interactions, the political intervention J is included in the coupling and subject to the inference. The coupling function *f_i_* is assumed to increase linearly and then saturates at a maximal value of 1, whereas *g_i_* shows the same behavior but remains restricted to the linear signal range. The strength and direction of the coupling is equivalent to the scale and sign of *γ_i_* and *c_i_* of the couplings *g_i_* and *f_i_*, respectively, subject to inference from empirical data using a stochastic model (see below). As we will not discuss group-specific mobility changes due to changes in PS variables, we drop the index of *γ_i_*, leading to the following closed loop equations:

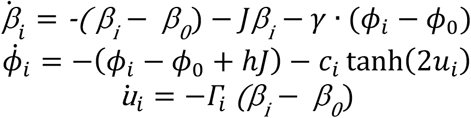

where *u_i_* = 0 for *J =* 0, and during the relaxation of restrictions *u_i_* is reset such that *u_i_*(*t*) *=* tanh^−1^(*u*(*t −* 1)(*J(t)/J*(*t −* l))^2^). Here Γ*_i_* <<1 reflects the time scale separation, that is both mobility via and affective state via *ϕ_i_* evolve on a time scale of the order of 1, whereas PS effects accumulate on a time scale of the order of *Γ_i_*. To include the impact of seasonality on the transmission ability of the virus (*12*), we adapt sinusoidal modulation of *β*_0_ with a period of one year and the peak being in Mid-January for the northern hemisphere. The seasonally varying *β*_0_(*t*) hence reads:

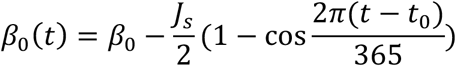

with*J_s_* being the magnitude of the variation that depends on the latitude and the climate. For the both Germany and Denmark, it is fixed at 40%, which is the level predicted for New York (*12*).

The statistical model was implemented in v2.22.1 of the Stan probabilistic programming language *(29)*, with parameters sampled using the No-U-Turn variant *(30)* of Hamiltonian Monte Carlo (31). The posterior draws were verified with the Stan ‘diagnose’ utility, which checks for transitions that hit the maximum tree depth, divergent transitions, low estimated Bayesian fraction of missing information (E-BFMI), low effective sample sizes and *Ȓ* values below 1.1 (indicating convergence of the MCMC method). Posterior distributions for key parameters and posterior predictive distributions for data were constructed, verifying that the data lie in a 95% confidence interval, to ensure model fit is reasonable.

